# Airway expression of SARS-CoV-2 receptor, ACE2, and proteases, TMPRSS2 and furin, in severe asthma

**DOI:** 10.1101/2020.06.29.20142091

**Authors:** Nazanin Zounemat Kermani, Woo-Jung Song, Alan Lunt, Yusef Badi, Ali Versi, Yike Guo, Kai Sun, Pank Bhavsar, Peter Howarth, Sven-Erik Dahlen, Peter J Sterk, Ratko Djukanovic, Ian M Adcock, Kian Fan Chung, on behalf the U-BIOPRED Consortium

## Abstract

**Background:** Patients with severe asthma may have a greater risk of dying from COVID-19 disease caused by SARS-CoV-2 virus. Angiotensin converting enzyme 2 (ACE2) receptor and enzyme proteases, transmembrane protease, serine 2 (TMPRSS2) and furin are needed for the attachment and invasion of the virus into host cells. We determined whether their expression in the airways of severe asthma patients is increased.

**Method:** We examined the microarray mRNA expression of ACE2, TMPRSS2 and furin in the sputum, bronchial brush and bronchial biopsies of participants in the European U-BIOPRED cohort.

**Results:** ACE2 and furin sputum gene expression was significantly increased in severe non-smoking asthma compared to mild-moderate asthma and healthy volunteers. By contrast, TMPRSS2 expression in bronchial biopsy and bronchial brushings was increased in severe smoking and ex-smoking asthmatics, and so was furin expression in bronchial brushings. Several clinical parameters including male gender, oral steroid use and nasal polyps were positively associated with ACE2, TMPRSS2 and furin expression levels. There was a higher expression of ACE2 and furin in the sputum neutrophilic molecular phenotype with inflammasome activation compared to the eosinophilic Type2-high or paucigranulocytic phenotypes. The enrichment score of the IL-13-Type2 gene signature was positively correlated with ACE2, TMPRSS2 and furin levels.

**Conclusion:** These key determinants of virus entry into the lungs may contribute to the poorer outcomes from COVID-19 disease in patients with severe asthma.

**“take home” message:** In severe asthma, gene expression of ACE, TMPRSS2 and furin are elevated compared to mild-moderate asthma and healthy volunteers, particularly in neutrophilic asthma. This might explain the increased risk of death in severe asthma afflicted with COVID19.

## Introduction

Coronavirus disease 2019 (COVID-19) is posing un unprecedented impact on global health. COVID-19 infection caused by the new SARS-CoV-2 virus has been reported to be more common or severe in patients with older age or pre-existing conditions (1-5). However, it is unclear whether asthma is associated with poor clinical outcomes in COVID-19. Although a few studies did not find a significant association between asthma and critical outcomes in COVID-19 patients (1-5), they did not characterize asthma in detail, including clinical and inflammatory phenotypes. A recent large population-based study in UK reported a higher risk of death in patients with severe asthma who developed COVID-19 disease (6), with another study indicating that those with chronic lung disease also had a poorer outcome (3). Thus, it is reasonable to speculate that asthma heterogeneity, particularly severity, is an important factor in understanding the risk of fatal outcomes in COVID-19 among asthmatic patients.

The spike protein of SARS-CoV-2 binds to the angiotensin converting enzyme 2 (ACE2) receptor allowing the virus to attach to the cell membrane of the host (7). The next step for entry into the cell depends on the spike protein being degraded by host cell enzyme proteases, such as transmembrane protease, serine 2 (TMPRSS2) and furin also known as PACE (<u>P</u>aired basic <u>A</u>mino acid <u>C</u>leaving <u>E</u>nzyme) (8, 9). Because this is the way by which SARS-CoV-2 infects and actively replicates in respiratory tract epithelial cells that expresses ACE2 receptors, we tested the hypothesis that the expression of these entry points for the virus may be different by asthma severity, and/or by clinical or molecular phenotypes.

We examined the transcriptomic data obtained from 3 airway compartment of airway cells, namely, bronchial brushings, bronchial biopsies and sputum-derived cells in patients with severe asthma from the U-BIOPRED (**U**nbiased **BIO**markers **P**redictive of **RE**spiratory **D**isease outcomes) cohort (10) and analysed the expression of genes that encode for ACE2, TMPRSS2 and furin. We also examined the relationship of the gene expression with co-morbidities that are known to be associated with high risk for COVID-19 and asthma molecular phenotypes or transcriptomic associated clusters (TACs), previously identified in U-BIOPRED.

## Methods

### Subjects

The European-wide U-BIOPRED cohort consisted of 4 groups of participants: Group A: severe non-smoking asthma (SAns), Group B: ever smokers with severe asthma (SAs), Group C: mild/moderate non-smoking asthmatics (MMA) and Group D: non-smoking healthy volunteers (HV) (10). Sputum cells, brushings of the lower airways, and bronchial biopsies (Supplementary Table S1) were obtained. Expression profiling was performed using U133 Plus 2.0 microarray (Affymetrix, Santa Clara, CA) on total RNA extracted and data assessed by multiarray average normalization. Demographics of the patients and healthy volunteers is shown in Supplementary Table S1. All U-BIOPRED participants gave signed informed consent to participate in the study which was approved by the local Ethics Committee of each country.

### Data analysis and statistics

Data were downloaded from the tranSMART platform (11). We analysed the gene expression of ACE2, TMPRSS2, and furin in the lower airway samples obtained from asthmatics and healthy subjects. A regression-based method (R package limma; version 3.45.0) was used to analyse these three genes with respect to the groups of interest, such as by the presence of asthma, across asthma severity subgroups, and also across 3 TACs (12). The presence of comorbidities (e.g. allergic rhinitis, nasal polyps, and eczema) was determined by physician diagnosis history. Age, sex, BMI (body mass index), and the use of oral corticosteroids (OCS) were adjusted for as covariates in the linear models. The Benjamini–Hochberg procedure controlled false discovery rate. Spearman’s rank correlation was used for correlation analysis.

### Signatures summarised by gene set variation analysis

Gene set variation analysis (GSVA) was used to calculate sample-wise enrichment scores (ESs) (13) for eight asthma-associated gene signatures. These gene sets each relate to a specific aspect of airway inflammation and asthma pathogenesis (Supplementary Table S2). The correlation between ESs and expression of ACE2, TMPRSS2, and furin was calculated using Spearman’s correlation. For group comparison *P* values were determined by differential expression analysis using moderated t-test, with adjustment for age, gender, body mass index and oral corticosteroid (R package limma; version 3.45.0).

## Results

### Intra-compartmental correlations of ACE2, TMPRSS2 and furin

ACE2, TMPRSS2 and furin mRNA expression was assessed in sputum (Fig 1A-C), bronchial brushing (Fig 1D-F), and bronchial biopsy (Fig 1G-I) compartments from HVs and asthmatics of different severity. There was no significant correlation between the expression of these three genes, in any compartment, except for a positive correlation between ACE2 and TMPRSS2 expression in sputum cells (n=89; r=0.252; p=0.010; Supplementary Table S3).

**Fig 1.**
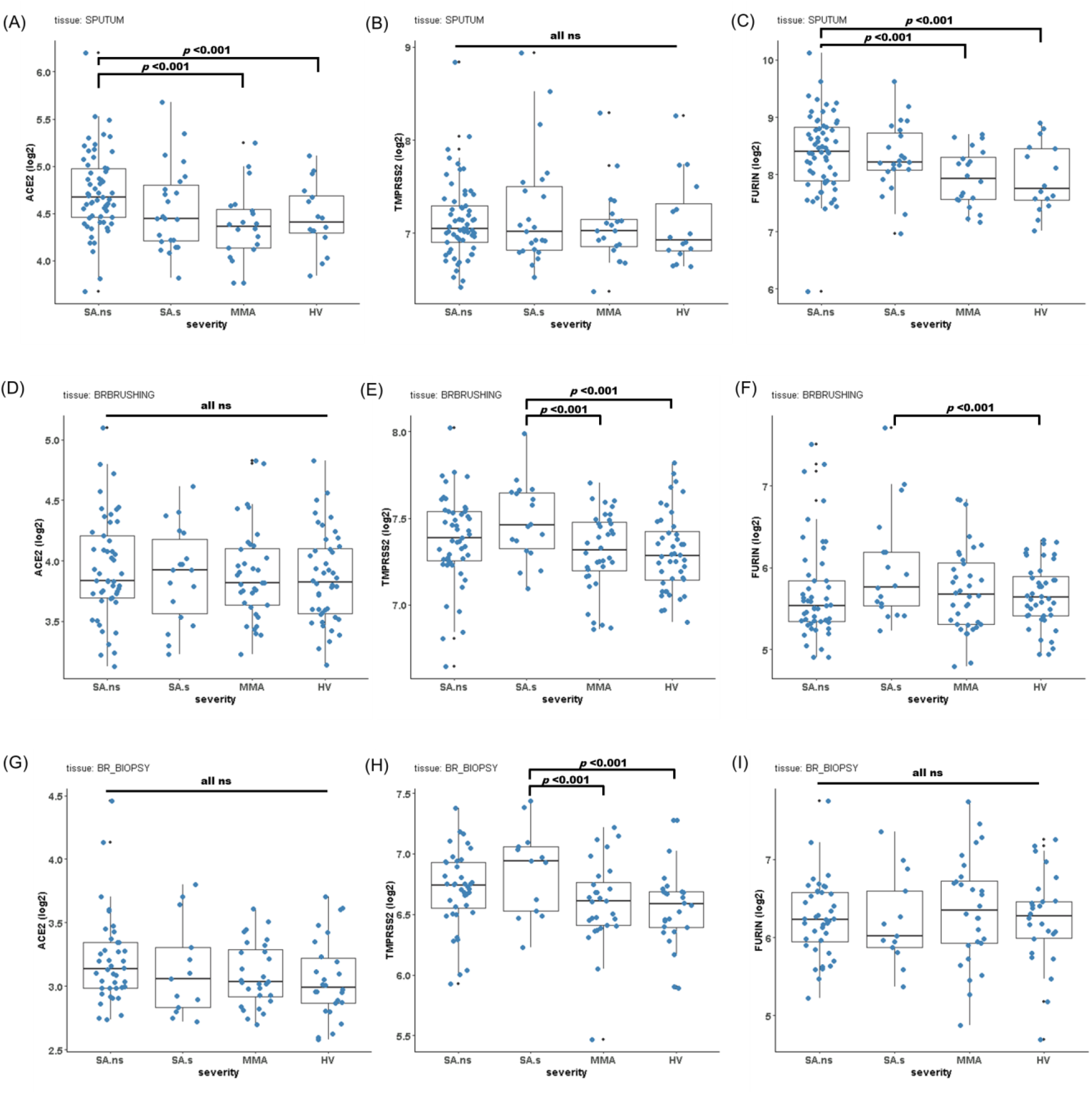
Comparison of ACE2, TMPRSS2 and furin gene expression levels (box-and-whisker plots showing median and interquartile range) between healthy volunteers and asthmatics in different airway compartments. ACE2 (A, D, G), TMPRSS2 (B, E, H) and furin (C, F, I) mRNA expression was assessed in (A) ACE2 in sputum (A-C), bronchial brushings (D-F), and bronchial biopsies (G-I) compartments from healthy volunteers and asthmatics of different severity. Abbreviations: SA.ns, severe asthmatics with no smoking history; SA.s, severe asthmatics with ever smoking history; MMA, mild-to-moderate asthmatics; HV, healthy volunteers; ns, not significant (*p* > 0.05). *P* values were determined by differential expression analysis: moderated t-test, with adjustment for age, gender, and oral corticosteroid use.

### Expression of ACE2

Overall, the gene expression levels were not significantly different between HVs and asthmatics when asthmatics analysed as a single group (data not shown). However, differences were revealed when sub-grouped by asthma severity. There was a significant increase in the expression of ACE2 in SAns versus MMA and HV in sputum, when adjusted for age, gender and OCS use (Fig 1A). Within asthmatics, the levels of ACE2 in sputum were higher in males and those on OCS (Table 1), and the expression of ACE2 in sputum positively correlated with the blood neutrophil count (r=0.21, p<0.05) and sputum neutrophil (%) count (r=0.28, p<0.01; Table 2). Expression levels of ACE2 mRNA were relatively lower in bronchial brushings and biopsies than in sputum (Supplementary Table S4). In bronchial brushings, ACE levels correlated with blood neutrophil count, but not with other baseline parameters (r=0.21, p<0.05; Table 2). In bronchial biopsies, the expression levels of ACE2 were higher in patients with nasal polyps but was not associated with other baseline parameters (Table 2).

**Table 1.** SARS-CoV-2 entry-related gene expression (Z-score) in the lower airways and their associations with baseline parameters within asthmatics.

| Parameters | Sputum (n=104) |  |  | Bronchial brushing (n=103) |  |  | Bronchial biopsy (n=81) |  |  |
| --- | --- | --- | --- | --- | --- | --- | --- | --- | --- |
|  | ACE2 | TMPRSS2 | Furin | ACE2 | TMPRSS2 | Furin | ACE2 | TMPRSS2 | Furin |
| Gender |  |  |  |  |  |  |  |  |  |
| Female | **-0.47 (-1.27 to 0.5)\*\*** | -0.45 (-1.89 to 1.6) | 0.08 (-0.54 to 0.8) | -0.33 (-1.02 to 0.7) | -0.17 (-1.35 to 1.1) | **-0.43 (-0.78 to 0.1)\*\*** | -0.22 (-0.68 to 0.8) | -0.35 (-1.33 to 1) | -0.06 (-0.67 to 0.6) |
| Male | **0.32 (-0.56 to 1.7)\*\*** | 0.01 (-1.57 to 1.4) | 0.05 (-0.88 to 0.6) | -0.06 (-0.88 to 1.1) | 0.61 (-0.73 to 1.7) | **0.08 (-0.49 to 0.98)\*\*** | -0.23 (-1.16 to 1.1) | 1.25 (-1.03 to 2.4) | -0.03 (-0.55 to 0.7) |
| Smoking |  |  |  |  |  |  |  |  |  |
| Never | 0.06 (-0.82 to 1.3) | -0.30 (-1.78 to 1) | 0.07 (-0.61 to 0.7) | -0.39 (-1.22 to 0.8) | -0.01 (-1.36 to 1.4) | -0.25 (-0.73 to 0.5) | -0.37 (-0.95 to 0.8) | 0.1 (-1.34 to 1.4) | -0.03 (-0.60 to 0.7) |
| Ever | -0.34 (-1.24 to 0.6) | -0.23 (-1.8 to 2) | 0.05 (-0.52 to 0.7) | -0.12 (-.54 to 1.1) | 0.74 (-0.74 to 2.1) | -0.18 (-0.59 to 0.9) | 0.07 (-1.13 to 1.1) | 1.7 (-1.02 to 3.1) | -0.23 (-0.65 to 0.6) |
| Nasal polyps |  |  |  |  |  |  |  |  |  |
| No | -0.50 (-1.2 to 1) | -0.40 (-1.83 to 1) | -0.02 (-0.78 to 0.6) | -0.01 (-1.04 to 1) | -0.004 (-1.0 to 1.2) | -0.32 (-0.74 to 0.4) | **-0.38 (-1.16 to 0.7)\*** | **0.09 (-1.39 to 1.3)\*** | -0.04 (-0.57 to 0.6) |
| Yes | 0.21 (-0.17 to 1.1) | 0.09 (-1.49 to 2.2) | 0.23 (-0.51 to 0.9) | -0.39 (-0.93 to 0.4) | 1.2 (-0.46 to 2.1) | -0.18 (-0.64 to 0.6) | **0.63 (-0.69 to 2)\*** | **1.61 (-0.05 to 2.7)\*** | -0.16 (-0.64 to 0.7) |
| Allergic rhinitis |  |  |  |  |  |  |  |  |  |
| No | -0.14 (-1.11 to 1) | -0.29 (-1.83 to 1.3) | 0.07 (-0.82 to 0.6) | -0.23 (-1.24 to 0.7) | 0.08 (-0.98 to 1.6) | -0.26 (-0.75 to 0.4) | -0.45 (-1.22 to 0.8) | **0.9 (-0.64 to 2.5)\*** | -0.03 (-0.57 to 0.8) |
| Yes | -0.08 (-0.64 to 1) | -0.04 (-1.71 to 1.8) | 0.07 (-0.42 to 0.9) | -0.27 (-0.87 to 1.1) | 0.38 (-0.90 to 1.5) | -0.26 (-0.67 to 0.5) | -0.15 (-0.77 to 0.9) | **0 (-1.9 to 1.4)\*** | -0.03 (-0.55 to 0.6) |
| Eczema |  |  |  |  |  |  |  |  |  |
| No | -0.07 (-0.88 to 1) | -0.04 (-1.90 to 1.8) | -0.03 (-0.82 to 0.6) | -0.38 (-1.23 to 0.8) | -0.09 (-0.97 to 1.2) | -0.22 (-0.7 to 0.6) | -0.38 (-0.99 to 0.7) | 0.3 (-1.08 to 2.3) | -0.04 (-0.7 to 0.6) |
| Yes | -0.30 (-1.27 to 1.3) | -0.98 (-1.81 to 1) | 0.22 (-0.29 to 1) | 0.32 (-0.8 to 1.1) | 0.64 (-0.90 to 1.6) | -0.28 (-0.75 to 0.3) | 0.02 (-1.01 to 1.1) | 0.3 (-1.33 to 1.9) | 0.003 (-0.52 to 0.6) |
| OCS use |  |  |  |  |  |  |  |  |  |
| No | **-0.48 (-1.17 to 0.6)\*** | -0.33 (-1.82 to 1.4) | 0.04 (-0.57 to 0.7) | -0.04 (-1.02 to 1.1) | -0.002 (-0.98 to 1.6) | -0.20 (-0.74 to 0.6) | 0.2 (-0.96 to 1.1) | 0.1 (-1.40 to 1.6) | -0.04 (-0.68 to 0.6) |
| Yes | **0.42 (-0.39 to 1.3)\*** | -0.04 (-1.64 to 2.2) | 0.08 (-0.62 to 0.8) | -0.45 (-0.95 to 0.3) | 1.06 (-0.06 to 1.9) | -0.27 (-0.68 to 0.9) | -0.6 (-1.11 to 0) | 0.5 (-0.73 to 2.3) | -0.06 (-0.39 to 0.7) |
| Atopy |  |  |  |  |  |  |  |  |  |
| No | -0.04 (-1.19 to 2) | -0.43 (-2.33 to 1.7) | 0.2 (-0.17 to 0.8) | **-0.74 (-1.1 to -0.13)\*** | 0.07 (-0.74 to 0.9) | -0.61 (-0.73 to 0.5) | -0.05 (-0.94 to 0.7) | 0.09 (-0.62 to 2.3) | 0.01 (-0.52 to 0.7) |
| Yes | -0.10 (-0.90 to 0.7) | -0.09 (-1.54 to 1.2) | 0 (-0.68 to 0.7) | **0.13 (-0.71 to 1.19)\*** | 0.37 (-1.07 to 1.7) | -0.16 (-0.66 to 0.6) | -0.37 (-1.04 to 1.1) | 0.23 (-1.49 to 1.6) | -0.09 (-0.70 to 0.6) |

Expression of ACE2, TMPRSS2, and Furin are described as median (interquartile range 25%-75%). The Chi-squared test was used for computing the *P* values. Statistically significant correlations are marked
bold and also indicated as follows: \**p* <0.05; \*\**p* <0.01; otherwise non-significant (*p* >0.05).

**Table 2.** Correlations between baseline parameters and ACE2, TMPRSS2 and Furin gene expression in asthmatics.

| Parameters | Sputum (n=104) |  |  | Bronchial brushing (n=103) |  |  | Bronchial biopsy (n=81) |  |  |
| --- | --- | --- | --- | --- | --- | --- | --- | --- | --- |
|  | ACE2 | TMPRSS2 | Furin | ACE2 | TMPRSS2 | Furin | ACE2 | TMPRSS2 | Furin |
| Age (years) | $r = 0.14$ | $r = -0.03$ | $r = 0.11$ | $r = -0.01$ | $r = 0.07$ | $r = 0.04$ | $r = 0.01$ | **$r = 0.23^*$** | $r = -0.16$ |
| BMI (kg/m<sup>2</sup>) | $r = 0.04$ | $r = 0.12$ | $r = 0.05$ | $r = 0.04$ | $r = 0.1$ | $r = -0.05$ | $r = -0.12$ | $r = 0.07$ | $r = -0.06$ |
| FEV1% predicted | $r = -0.17$ | $r = -0.04$ | **$r = -0.21^*$** | $r = -0.06$ | $r = -0.14$ | $r = -0.19$ | $r = -0.08$ | **$r = -0.23^*$** | $r = 0.04$ |
| FeNO (ppb) | $r = 0.06$ | $r = 0.01$ | **$r = -0.26^*$** | $r = 0.11$ | $r = 0.05$ | $r = -0.04$ | $r = -0.01$ | $r = -0.1$ | $r = -0.04$ |
| Total IgE (IU/mL) | $r = 0.14$ | $r = 0$ | $r = 0.02$ | $r = 0.18$ | $r = 0.19$ | $r = 0.02$ | $r = 0.15$ | $r = 0$ | $r = -0.12$ |
| Serum periostin (ng/mL) | $r = 0.08$ | $r = 0$ | $r = 0.08$ | $r = -0.09$ | **$r = -0.24^*$** | $r = 0.01$ | $r = 0.06$ | $r = -0.02$ | $r = -0.16$ |
| Blood eosinophils (x 10<sup>9</sup>/L) | $r = 0.18$ | $r = 0.03$ | $r = 0.02$ | $r = 0.07$ | $r = -0.02$ | $r = 0.13$ | $r = -0.07$ | $r = -0.01$ | $r = -0.03$ |
| Blood neutrophils (x 10<sup>9</sup>/L) | **$r = 0.21^*$** | $r = 0.06$ | **$r = 0.24^*$** | **$r = 0.21^*$** | **$r = 0.21^*$** | $r = 0.17$ | $r = 0.15$ | **$r = 0.34^{**}$** | $r = 0.15$ |
| Sputum eosinophil % | $r = 0.07$ | $r = -0.03$ | $r = -0.1$ | $r = -0.11$ (n=43) | $r = 0.01$ (n=43) | $r = -0.12$ (n=43) | $r = 0.03$ (n=35) | $r = 0$ (n=35) | $r = 0.1$ (n=35) |
| Sputum neutrophil % | **$r = 0.28^{**}$** | $r = 0.13$ | **$r = 0.6^{***}$** | $r = 0.07$ (n=43) | $r = 0$ (n=43) | $r = 0.24$ (n=43) | $r = 0.26$ (n=35) | $r = -0.03$ (n=35) | $r = -0.06$ (n=35) |

Abbreviations: BMI, body mass index; FEV1, forced expiratory volume in 1 second; FeNO, exhaled nitric oxide fraction.
Association of ACE2, TMPRSS2, and furin expression with numerical variables were measured and tested using Spearman's rank-order correlation. Statistically significant correlations are marked bold and also indicated as follows: \* $p < 0.05$ ; \*\* $p < 0.01$ ; \*\*\* $p < 0.001$ . Otherwise non-significant ( $p > 0.05$ ).

### Expression of TMPRSS2

TMPRSS2 mRNA expression levels were higher in bronchial brushings than in sputum or bronchial biopsies (Supplementary Table S4). In bronchial brushings, there was a significantly increased expression of TMPRSS2 in SAs (versus MMA and HV) (Figure 1E), and the expression levels of TMPRSS2 positively correlated with blood neutrophils (r=0.21, p<0.05) (Table 2). In bronchial biopsies, TMPRSS2 levels were significantly higher in SAs (versus MMA and HV) (Figure 1H) and were increased in subjects with nasal polyps but also in those without allergic rhinitis (Table 1). However, TMPRSS2 expression was similar across all groups in sputum (Fig 1B).

### Expression of furin

Expression levels of furin were higher in sputum than in bronchial brushings and biopsies (Supplementary Table S4). In sputum, expression of furin was significantly greater in SAns compared to MMA and HVs (Fig 1C). Within the whole asthmatic group, sputum furin levels negatively correlated with FEV1 (% of predicted) (r=-0.21, p<0.05) and FeNO levels (r=-0.26, p<0.05), but positively with blood neutrophil count (r=0.24, p<0.05) and stronger with sputum neutrophil (%) (r=0.6, p<0.001) (Table 2). Meanwhile, in bronchial brushings, furin level was significantly higher in SAs than HVs (Figure 1F); however, it did not correlate with clinical parameters (Tables 1 and 2). Furin levels in bronchial biopsy were not significantly different across groups or by clinical parameters (Figure 1I and Tables 1 and 2).

### Expression of ACE2, TMPRSS2 and furin in molecular phenotypes of asthma

We determined the expression of these three genes in sputum amongst the three previously identified TACs (12, 14). As previously described, analysis of sputum transcriptomics in U-BIOPRED produced three molecular endotypes (12). The expression of ACE2 was highest in TAC2, an endotype characterized by sputum neutrophilic inflammation and inflammasome activation signature, compared to TAC1 (eosinophilic inflammation, T2-high) and TAC3 (paucigranulocytic inflammation), with levels higher than in HVs (Fig 2A). Furin expression showed a similar trend as that of ACE2 with the highest expression in TAC2, significantly higher than in TAC1 (Fig 2C). There was no significant difference in TMPRSS2 expression among the three TACs (Fig 2B).

**Fig 2.**
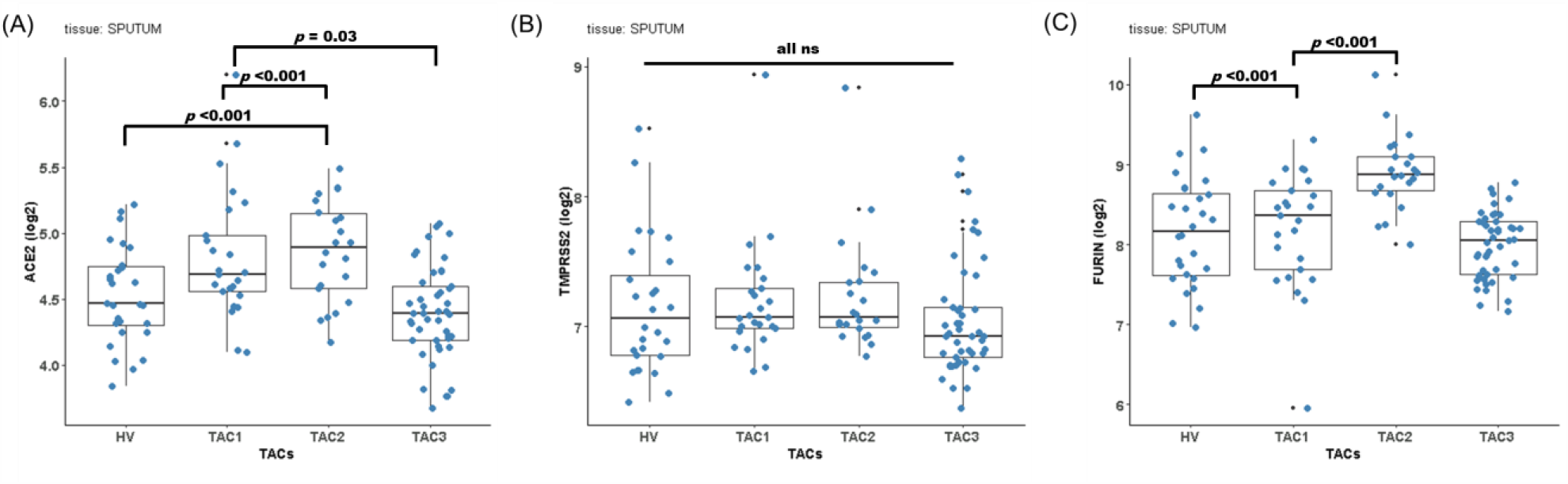
Comparison of (A) ACE2, (B) TMPRSS2 and (C) furin gene expression levels across transcriptomic-associated clusters in sputum samples. Data shown as box-and-whisker plots showing median and interquartile range. Abbreviations: HV, healthy volunteers; TAC, transcriptomic-associated cluster; ns, not significant (*p* > 0.05). *P* values were determined by differential expression analysis: moderated t-test, with adjustment for age, gender, body mass index and oral corticosteroid use.

**Fig 3.**
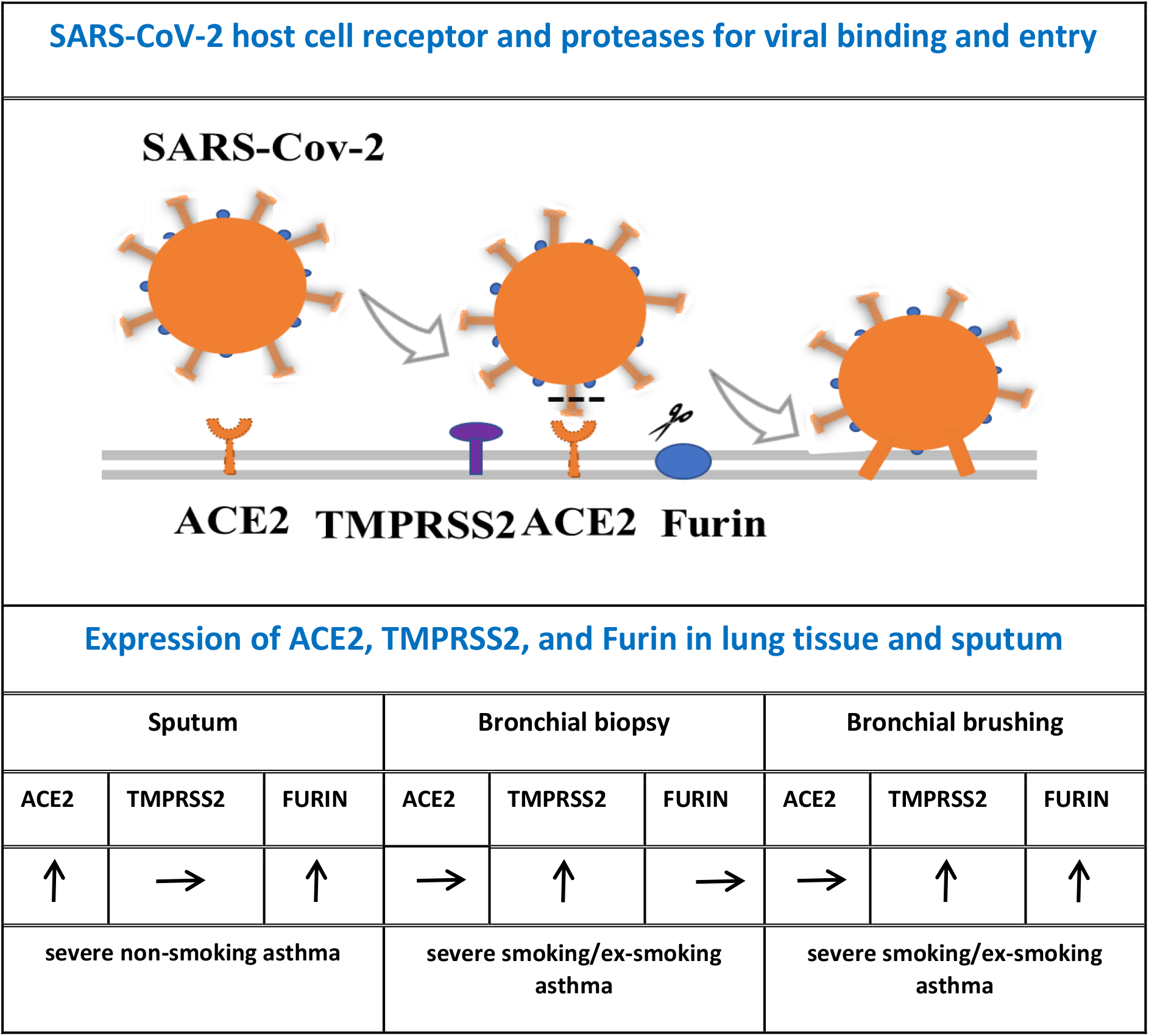
**Upper panel:** Interaction of the spike proteins of SARS-CoV-2 virus with the ACE2 receptor on host cell membrane, followed by the proteases, furin and TMPRSS2, on the exterior of the host cell breaks the spike protein at one or more cleavage sites. Fusion peptides are exposed fusing with the viral membrane with the host membrane. **Lower panel**: Expression levels of ACE2, TMPRSS2 and furin in sputum, bronchial brushings and bronchial biopsies of patients with severe non-smoking asthma and severe smoking/ex-smoking asthma compared to healthy volunteers.

### Gene signatures and expression of ACE2, TMPRSS2 and furin

To further determine the relationship between gene signatures of interest and the expression levels of ACE2, TMPRSS2 and furin, we measured by GSVA the ESs of IL-13-Th2, eosinophil activation, IL-17, neutrophil activation, IL-6-trans-signalling (IL-6-TS), and inflammasome signatures (Supplementary Table S2). The ES of IL-13-Th2 signature showed positive correlations with ACE2, TMPRSS2 and furin levels in the majority of compartments analysed (Table 3). The ES of eosinophil activation weakly correlated weakly with ACE2 in sputum (r=0.18, p<0.05), and with furin in bronchial biopsies (r=0.26, p<0.05). However, furin levels in all 3 compartments correlated with the neutrophil activation and IL-6-TS signatures with the highest correlation in sputum (r=0.51, p<0.001; and r=0.54, p<0.001, respectively). Inflammasome activation also significantly correlated with furin levels in sputum (r=0.49, p<0.001) and in bronchial brushings (r=0.30, p<0.001).

**Table 3.**
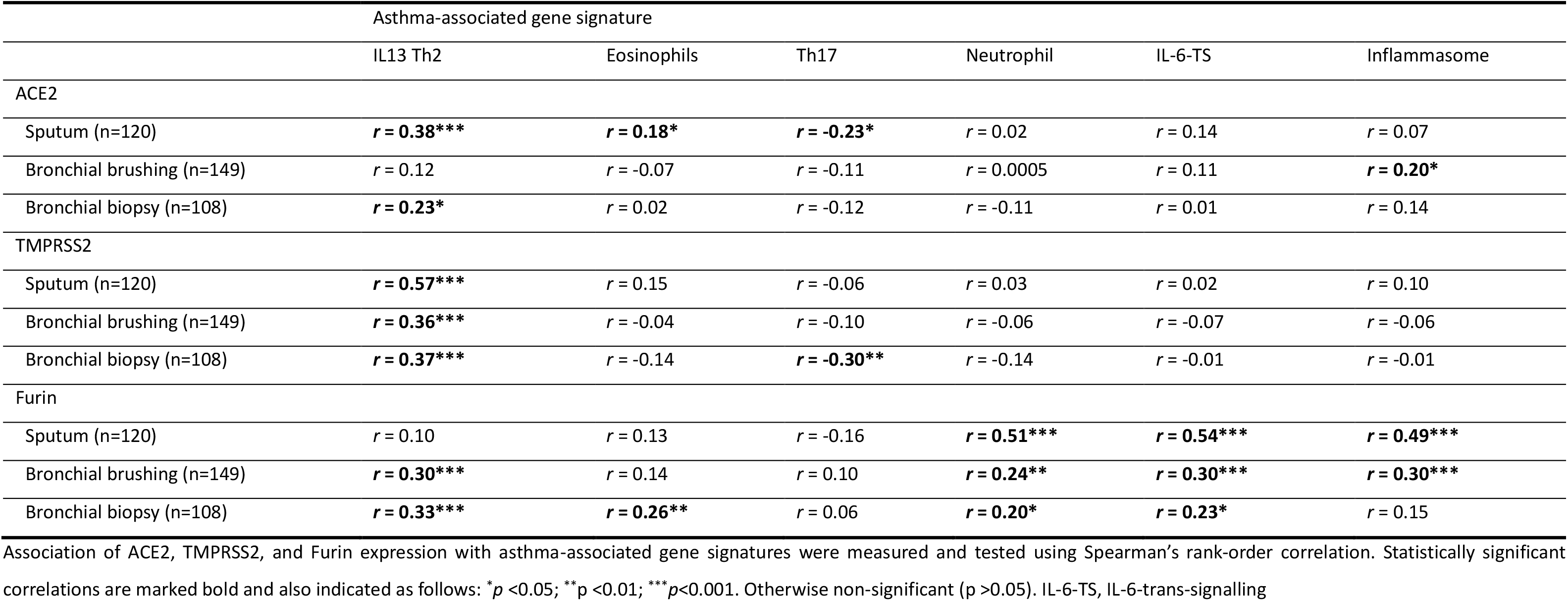
Summary of correlations between SARS-CoV-2 entry-related gene expression and asthma-associated gene signatures.

## Discussion

COVID-19 is a disease that spreads easily, hence the pandemic, causes severe illness in around 20% of affected individuals, with up to 33% deaths in those admitted to hospital. The factors that determine the clinical picture are slowly becoming apparent, but elucidation of the molecular mechanisms that must be the defining factors for all clinical disease has not been achieved, especially in the context of individual chronic diseases, like asthma, and the various phenotypes of this condition that varies from mild to very severe. Analysing three cell compartments (biopsies, brushings, and sputum cells) this study provides compelling evidence for over expression of the factors that may determine the entry of the SARS-CoV2 into cells, ACE2, TMPRSS2 and furin in severe asthma. In contrast, their expression in mild to moderate asthmatics compared to mild-moderate asthma was similar to healthy controls. Together, these observations suggest that patients with severe asthma, and not those with mild disease, have the pathobiological substrate that determines the strength of infection by SARS-CoV-2. Our findings also suggest that the associations between SARS-CoV2 infection and asthma are likely to be complex, and support the epidemiological observation (6) that severe asthma may increase the risk of severe morbidity or mortality if infected with SARS-CoV-2 virus with increased probability in males, OCS users or those with comorbid nasal polyps.

In sputum, ACE2 and furin were most highly expressed in severe non-smoking asthma (SAns), while in bronchial biopsies and bronchial brushings, TMPRSS2 was more highly expressed in severe smoking/ex-smoking asthma (SAs). Further analysis suggested that several factors including male gender, OCS use or comorbid nasal polyps have a higher expression of these genes in sputum, bronchial biopsies or bronchial brushings. We also found that a specific molecular cluster (TAC2) in asthma, characterized by predominant sputum neutrophilic inflammation and inflammasome activation (12), had a higher expression of ACE2 and furin, than the other TACs. It was followed by TAC1, a predominantly sputum eosinophilic phenotype with the highest enrichment of gene signatures for IL-13/Type-2 (T2) inflammation, and lowest in TAC3, with a paucigranulocytic sputum associated with mild eosinophilia and increased metabolic and mitochondrial function genes. However, the expression of TMPRSS2 was not different amongst the 3 TACs or compared to healthy individuals. These findings are supported by the significant correlations between ACE2 and furin expression with gene signatures of relevance to asthma, such as neutrophil activation, IL-6-TS, inflammasome, or IL13-Th2 pathways. Thus, there is the possibility that these pathways may increase the expression of ACE2 or furin. Collectively, these suggest that molecular heterogeneity may be associated with the varying risk of severe COVID-19 outcomes in asthmatics.

Olfactory and taste disturbance is a common symptom of COVID-19, and the nasal epithelium is suggested to be a major route of viral infection (15, 16). In this study, we did not report nasal brushing data in detail, as three gene expression levels in nasal brushings did not differ by asthma severity or molecular phenotype (data not presented) and also were not significantly different in those with nasal polyps (Supplementary Fig S1). However, the absolute gene expression levels were higher in nasal brushings than in other airway compartments (Supplementary Table S4), supporting a recent expression analysis report (16). Based on the expression patterns in the upper and lower airways, we suggest that severe asthmatics might have a higher risk of poorer outcomes from COVID-19, although the risk of infection via the nasal route is unlikely to be different.

A strength of our analysis is that it evaluates expression in a range of airway cell populations (epithelial brushings, bronchial biopsies and sputum cells) in well-characterised subjects allowing, particularly in severe asthma, assessment in different sub-divided, phenotypic populations. Like a recent publication that found no difference in gene expression of ACE2, TMPRSS2 and furin between healthy, mild-to-moderate asthma and severe asthma in bronchial brush samples, we did not find differences when the severe asthmatics were those classified as non-smokers but did find differences in those with severe asthma who were either current smokers or ex-smokers with a 5-pack-year history or more. By contrast it was only the severe non-smoking asthmatics that had increased gene expression of ACE2 and TMPRSS2 in sputum, a difference that was not evident when mild/moderate and severe asthma were all included as a mixed asthma population in a previous study (17).

Consistent with the importance of focusing on phenotypic populations, in asthmatic children with respiratory allergy and allergen exposure have reduced levels of ACE2 expression in nasal/bronchial epithelial brushings compared to those without allergies (18). Similarly, in bronchial brushings in adults with mild-moderate asthma, ACE2 expression has been reported to be significantly reduced in those defined as T2-high asthma (high expression of three-gene signature; CLCA1, POSTN, SERPINB2) compared to those with T2-low asthma (19). Our failure to confirm these findings in severe asthma may be due to confounding by therapy, the nature of the underlying type 2 inflammatory process (Th2 allergen driven vs ILC2 non-allergen driven) as well as the analytic approach. Using GSVA, we showed that the expression of the T2 signature to correlate positively with the expression of the 3 genes in most of the 3 compartments, while the neutrophil activation signature was only correlated with furin expression. These differences may reflect the complexity of asthma driving mechanisms whereby the presence of differing percentages of Th2, Th17 and Th1 pathways in each asthmatic individual (20, 21) may effect upon the overall expression of the SARS-CoV-2 entry and activation genes.

Another strength of our study is the large sample size and the fact that the U-BIOPRED cohort came from one single large study that recruited healthy controls, mild-moderate asthmatics and severe asthmatics using the same pre-defined protocol. In addition, we compared the expression of these three pivotal genes for SARS-CoV-2 virus entry in three different compartments of relevance: bronchial epithelial cells from brushings, bronchial biopsy with epithelial and submucosal cells and sputum cells with granulocyte inflammation. It is this uniform approach that gives confidence in the observation that the levels of ACE2, TRPMSS2 and furin are higher in severe asthma compared to mild moderate asthma, particularly in males, with nasal polyps or on oral corticosteroid therapy, and those with an eosinophilic and neutrophilic inflammation. However, a limitation of our study is that these data cannot apportion cause and effect as this is a comparative study with correlation analyses. In addition, we do not know whether the increased gene expression is associated with greater protein expression. Further studies are therefore warranted.

## Data Availability

The data will be available to any requests.

## Acknowledgements

U-BIOPRED was supported by an Innovative Medicines Initiative Joint Undertaking (No.115010), resources from the European Union’s Seventh Framework Programme (FP7/2007-2013) and EFPIA companies’ in-kind contribution (www.imi.europa.eu). We thank the U-BIOPRED Project team for their contribution.

## Notes

### Competing Interest Statement

KFC has received honoraria for participating in Advisory Board meetings of GSK, AZ, Roche, Novartis, Merck, BI, TEVA and Shionogi regarding treatments for asthma, chronic obstructive pulmonary disease and chronic cough and has also been renumerated for speaking engagements. Dr. Adcock has nothing to disclose.Zounemat Kermani has nothing to disclose.Dr. Bhavsar has nothing to disclose.Dr. Song has nothing to disclose.Dr. Sterk has nothing to disclose.Dr. Sterk reports grants from public-private funding by the Innovative Medicines Innitiative (IMI) covered by the European Union (EU) and the European Federation of Pharmaceutical Industries and Associations (EFPIA), during the conduct of the study; .Dr. Dahlen reports personal fees from AZ, Cayman Chemical, GSK, Novartis, Sanofi, Regeneron, TEVA, outside the submitted work; .Dr Djukanovic reports receiving fees for lectures at symposia organised by Novartis, AstraZeneca and TEVA, consultation for TEVA and Novartis as member of advisory boards, and participation in a scientific discussion about asthma organised by GlaxoSmithKline. He is a co-founder and current consultant, and has shares in Synairgen, a University of Southampton spin out company.Dr Kai Sun, Alan Lunt, Peter Howarth and Yike Guo have nothing to disclose.

### Clinical Trial

Not applicable

### Author Declarations

This was provided by the local Institutution of each participating country

